# A randomized controlled trial comparing epinephrine and dexamethasone to placebo in the treatment of infants with bronchiolitis (BIPED Study): A Statistical Analysis Plan

**DOI:** 10.1101/2025.06.03.25328902

**Authors:** Anna Heath, David Rios, Kristina I Vogel, Tremaine Rowe, Natasha Wills-Ibarra, Ed Oakley, Martin Offringa, Petros Pechlivanoglou, Terry P Klassen, Stuart R Dalziel, Amy C Plint, the KidsCan Pediatric Emergency Research Canada (PERC) Innovative Pediatric Clinical Trials and Pediatric Research in Emergency Department International Collaborative (PREDICT) BIPED Study Team

## Abstract

**Background:** Bronchiolitis is a common lung infection that affects infants and young children. While most children can be treated at home, some require hospitalisation where supportive care, such as fluids and oxygen, is the suggested treatment. Bronchiolitis is the leading cause of infant hospitalisation in developed countries and exerts a significant burden on the healthcare system. The aim of the Bronchiolitis in Infants Placebo Versus Epinephrine and Dexamethasone (BIPED) study is to evaluate the effects of a combination of epinephrine and dexamethasone, given during initial presentation at the emergency department, on hospitalisation for bronchiolitis. This article outlines the statistical analysis plan (SAP) for the BIPED study.

**Methods/Design:** The BIPED study is a Phase III, multi-centre, randomised, controlled, double-blinded superiority, placebo-controlled trial to determine whether the combination of epinephrine and dexamethasone is successful in reducing hospitalisation for bronchiolitis up to seven days following presentation at an emergency department with bronchiolitis. The secondary outcomes include hospital admissions for bronchiolitis at the emergency department enrolment visit, and all cause hospital admissions, health care provider visits and health care-related costs in the 21 days following enrolment. The safety outcomes are gastrointestinal bleeding, serious bacterial infection, severe varicella and death.

**Discussion:** The BIPED study will provide evidence on whether a combination of epinephrine and dexamethasone reduces hospitalisation in infants following presentation to the emergency department with bronchiolitis. These data will be analyzed using this SAP, submitted before the data became available for analysis, to reduce the risk of bias in our reported outcomes.

**Trial Registration:** ClinicalTrials.gov Identifier: NCT03567473

## Background

Bronchiolitis exerts substantial pressure on healthcare systems in developed countries, both in terms of health-related expenditure and hospitalisation, and is the primary cause of hospitalisations in infants [1, 2, 3, 4]. Despite this burden and significant research expenditure, there is no evidence on an effective treatment for bronchiolitis [5]. International guidelines recommend only supportive care, including the administration of fluids and oxygen as necessary [6]. However, a large multicenter trial indicated that a combination of nebulised epinephrine and oral dexamethasone could potentially reduce hospitalisations for children with bronchiolitis, compared to placebo [7]. Furthermore, international guidelines have called for research to determine whether this combination therapy is an effective treatment for bronchiolitis [8, 9, 6]. The Bronchiolitis in Infants Placebo Versus Epinephrine and Dexamethasone (BIPED) study aims to collect this evidence.

The BIPED study is a phase III, multi-centre, randomised, controlled, double blinded, superiority placebo-controlled trial that will investigate the combination of two treatments of inhaled epinephrine and two doses of oral dexamethasone as a treatment for bronchiolitis. The BIPED study will enrol infants who present at the emergency department (ED) with bronchiolitis. The study protocol has been published separately [10]. This article outlines the final statistical analysis plan (SAP) for the BIPED study. This SAP has been published before completing the data lock for the BIPED study and aims to reduce bias in the final trial report.

## Objectives

The primary aim of the BIPED study is to determine whether infants with bronchiolitis treated with inhaled epinephrine in the ED and a two-day course of oral dexamethasone will have fewer hospitalisations related to bronchiolitis over the seven days following ED presentation, compared to infants treated with placebo. We will enrol infants aged between 60 days and 12 months who present at the ED with bronchiolitis during the period deemed to be the season for respiratory syncytial virus (RSV) bronchiolitis. The secondary objectives are to investigate the effect of the combination of epinephrine and dexamethasone on hospital admission for bronchiolitis at the enrolment ED visit, all cause hospital admissions at 21 days, and health care provider visits (including ED visits) and health care related costs within 21 days post-enrollment in the trial. The primary outcome will also be investigated in the participants who present at the ED with their first episode of bronchiolitis.

## Method/Design

### Design and setting

The BIPED study is a phase III study conducted in 12 sites across Canada, Australia and New Zealand. All participating hospitals are members of the Pediatric Emergency Research Canada (PERC) network [11] or the Paediatric Research in Emergency Departments International Collaboration (PREDICT) [12], both of whom are affiliated with the Pediatric Emergency Research Networks (PERN). The BIPED study will define bronchiolitis as an episode of wheezing or crackles in an infant aged between 60 days and 12 months, alongside signs of an upper respiratory tract infection during the period deemed to be the season for RSV bronchiolitis. To reflect international clinical guidelines and practice, we have not restricted our definition of bronchiolitis to the first episode of wheezing or crackles and will enrol infants with second and subsequent episodes of bronchiolitis, with each infant only enrolled once into the study.

The active treatment in the BIPED study is two treatments of epinephrine (either via nebulisation (3 mg) or via metered dose inhaler and spacer (MDI-s) (625 mcg)) given 30 minutes apart in the ED and two doses of once daily oral dexamethasone (0.6 mg/kg per dose, up to a maximum of 10mg), known as EpiDex. The first dose of dexamethasone will be given immediately before the first dose of epinephrine and the second dose will be given 24 hours later. The BIPED study will compare EpiDex to a placebo control (oral dexamethasone vehicle and nebulised normal saline or an inert placebo by MDI-s) by randomising participants in a 1:1 ratio. All study personnel, bedside clinicians, and participants/caregivers are blinded to the study group assignment. Participants will be followed up for 21 days following their ED enrolment visit with the primary outcome collected at seven days (168 hours) post enrolment.

### Study Protocol Development and Conduct

The BIPED study was registered on ClinicalTrials.gov on June 25, 2018, registration number NCT03567473. The BIPED study protocol received approval from Health Canada and local research ethics boards (REB) at each site prior to commencing enrolment. Informed consent will be obtained from all legal guardians before administering the study interventions. The BIPED study is being undertaken within the Canadian KidsCAN-PERC iPCT network [13] and the Australian-New Zealand PREDICT network. The data management and trial oversight, including safety, is being undertaken by the KidsCan-PERC iPCT trials network through a centralised structure and independent data and safety monitoring board (DSMB).

### Randomisation and data collection

Eligible participants will be randomised using permuted-block randomisation, stratified by site. This randomisation strategy ensures that differences in standard practice across sites will be equally represented in each study group. Additionally, block randomisation ensures that seasonal variation in the response to therapy will be comparable across the two groups. The statistician performing providing the randomisation lists is not involved in the study in another capacity.

Study participant research data will be stored using REDCap electronic data capture system [14], based at a secure data coordination centre (DCC) at Women and Children’s Health Research Institute (WCHRI), University of Alberta [15]. Individual participants will be identified with a unique study identification number.

### Primary Outcome

The primary outcome is admission to hospital for bronchiolitis within seven days (168 hours) of study enrolment. BIPED study defines hospitalisation as any of the following conditions:

1. The participant is admitted to an inpatient ward.
2. An ED length of stay of 12 hours or greater.
3. A combined ED and observation unit length of stay of 12 hours or greater.

ED length of stay at enrollment visit is defined as the time between the administration of first study medication (or time of consent when no study medications are received) and discharge from the ED or observation unit. The length of stay for all return ED visits will be measured from time of triage or registration, whichever is sooner.

### Secondary outcomes

#### Efficacy Outcomes

The BIPED study will include four secondary outcomes:

1. Admission to hospital for bronchiolitis at enrolment ED visit.
2. All-cause hospital admission within 21 days following enrollment ED visit.
3. All-cause health care provider visits (including ED visits) within 21 days following enrollment ED visit.
4. Health care related costs within 21 days of enrollment following enrollment ED visit.

#### Safety Outcomes

The safety of the EpiDex combination will be assessed with the following outcomes:

1. Gastrointestinal bleeding (involving melena or frank blood per rectum not attributed to other causes by the treating physician).
2. Serious bacterial infection (defined as meningitis, osteomyelitis or septicaemia).
3. Severe varicella (defined in the protocol [10]).
4. Death.

#### Exploratory Outcomes

The BIPED study will collect 10 additional exploratory outcomes:

a. Admission to hospital for bronchiolitis within 21 days following enrollment ED visit.
b. Intensive care unit (ICU) admissions within 21 days following enrollment ED visit for bronchiolitis and requiring intubation or continuous positive airway pressure (CPAP).
c. All cause hospital admission within seven days following enrollment ED visit.
d. All cause ED visits within 21 days of enrollment ED visit.
e. Length of stay (in hours) for the enrollment ED visit for participants who are discharged at the enrollment ED visit.
f. Length of hospital admission (in hours) for participants admitted at their enrollment visit.
g. Resolution of symptoms as documented on a standardised questionnaire during the telephone or email at day 7 and 21 days.
h. Out of pocket expenses borne by the family/caregiver in the 21 days following enrollment ED visit.
i. Health care utilization for respiratory illness up to 18 years of age
j. Development of respiratory illnesses such as asthma and recurrent wheezing up to 18 years of age.

### Sample Size Calculation

The sample size was determined using the average length criterion (ALC) [16], a Bayesian method for sample size determination. Using the ALC, we chose the smallest sample size for which the average length of the 95% high density posterior credible interval for the difference in admission rates between placebo and EpiDex is below 9%. We determined the average length of the posterior credible interval across the prior-predictive distribution for the data [48]. To determine the relevant prior distribution, we performed an expert elicitation for the probability of hospitalisation under placebo and EpiDex [17]. We simulated 1500 studies from the prior-predictive distribution and computed the posterior distribution for the absolute difference using 5000 simulations to estimate the average length of the 95% posterior credible intervals. Based on this simulation, the sample size of 410 participants per arm for the BIPED study was selected [17]. We allowed for a 5% loss to follow up, leading to a total target recruitment of 864 participants, 432 per arm.

### Interim analysis and stopping guidance

There are no planned interim analyses of efficacy outcomes. The DSMB will review all safety outcomes at least once a year for the duration of the study. The DSMB, alongside the trial steering committee, will consider stopping for safety concerns.

### Statistical Analysis Plan

#### Statistical Principals

Outcomes will be collected with a standardised survey issued to parents/caregivers either over email or by telephone seven and 21 days after enrollment in the BIPED study. Non-responders will be sent a second email and, if necessary, up to six attempted phone calls over three days. Once 21 days have elapsed following enrollment, the participant’s medical record will be reviewed to track any return ED visits, hospital admissions and additional healthcare contact. Admissions and ED visits reported by caregivers to study hospitals must be confirmed on medical record review. Similarly, at the six study hospitals where admissions and ED visits to non-study hospitals can be confirmed by administrative database review, non-study hospital ED visits and admissions reported on the survey will be confirmed. For study sites where visits and admissions to non-study hospitals cannot be confirmed by administrative database review, the caregiver’s report of and reason for these visits/admissions will be used to determine whether the participant met the study definition of admission. The final trial analysis will take place after every participant has reached 21 days after enrollment and all the data have been collected and cleaned. The analysis will be performed in a blinded manner.

The final trial analysis will use a Bayesian inferential framework based on the posterior probability that the EpiDex combination is superior to placebo, for the primary and secondary outcomes. It will follow the intention-to-treat principle. The EpiDex combination will be declared superior to placebo if the posterior probability of superiority is above 99%, with simulations confirming a Type 1 error rate of 4.1% with a probability of hospitalisation equal to 0.35 and a power of 81% with a target difference of 8%. Due to the likelihood principle, corrections for multiple comparisons will not be used. All estimates for the treatment effect will be reported using 95% high density posterior credible intervals, calculated using R [18] interfacing with Bayesian software such as JAGS, stan or INLA [19, 20, 21].

#### Handling of missing data

Participant medical record/database review will minimise the presence of missing data for many outcomes. Descriptive statistics on the proportion of missing data by treatment arm will be provided for the outcomes and the covariates. If the level of missingness is below 5%, we will undertake a complete case analysis. If greater than 5% of the data are missing, we will evaluate if there are any differences between the baseline characteristics for participants with and without missing data. Using this evidence, we will determine whether a complete case analysis is valid. If covariate outcomes are missing and a complete case analysis is not valid, the primary analysis will be a joint Bayesian analysis of the missingness mechanism and outcome model. In general, we will not jointly model or impute missing outcomes. If outcomes are missing and a complete case analysis is not used, then we will report descriptive statistics for this outcome and the reasons that individuals were lost to follow-up.

#### Patient Flow

We will use a CONSORT 2010 flow diagram to present the participant flow [22]. This diagram will report how many infants were screened, eligible for the trial and excluded due to our exclusion criteria. We will record the age, sex for all screened patients, and provide descriptive statistics for these quantities. The CONSORT diagram will also detail the number of participants who are lost to follow-up and who discontinue the intervention. Finally, we will also highlight the number of participants who were randomised but excluded from the primary analysis.

Participants will be free to withdraw from the BIPED study at any time and an investigator can discontinue or withdraw a patient if it would not be in the best interest of the participant to continue. The reason for, and the number of, study withdrawals will be tabulated for each treatment option.

#### Protocol Deviations

We define protocol deviations as participants who are randomised to either treatment but do not receive all of the study medications in the ED and participants who receive their ED medications at the incorrect dose and time, allowing for 5% tolerance on dose. Treatment adherence is taking all the trial medications. Adherence will be monitored by research staff in the ED and the additional dose of dexamethasone will be assessed through questions on the day seven follow-up survey. We will report the proportion of patients who had full adherence by treatment group. The number (and percentage) of patients with protocol deviations will be summarised by treatment group. The number of patients included in the intention-to-treat analysis will be used to calculate the percentage of participants with a protocol deviation. We will not undertake inferential statistical analysis for protocol deviations.

#### Analysis Populations

The primary analysis will include all randomised participants for whom outcome data are available and who do not withdraw consent for the use of their data as an intention-to-treat analysis. A per protocol analysis that includes only eligible participants for whom we have outcome data, who do not withdraw consent for the use of their data, who complete all their ED based medications and receive the correct dose (within +/- 5%) and timing of medications will also be performed. Finally, we will examine our outcomes in the per protocol population while also excluding participants who did not take their second dose of oral medication (unless admitted to hospital within the 40-hour second dose of oral medication window).

#### Baseline Characteristics

We will collect baseline information about the participants’ age, sex, weight, race, indigenous status and home environment, which includes the presence of smokers and attendance at daycare. We will also collect baseline clinical characteristics (respiratory distress assessment index (RDAI) [23], oxygen saturation, respiratory rate, heart rate, and temperature), the length of symptoms and treatments given prior to initial ED presentation, the medical history of the participant (such as breastfeeding, maternal smoking during pregnancy, prematurity, previous intubations, personal history of atopy [environmental allergy, food allergy and eczema, and previous significant/chronic illness], and previous episodes of wheezing/bronchiolitis) and family history of atopy (including asthma). Finally, if respiratory virus testing is available, the virus status will be reported.

Baseline characteristics will be compared across treatment group and reported based on their data type. Specifically, categorical data will be summarised by numbers of participants within each category and percentages. If the continuous data are sufficiently normal, they will be summarised with the mean and standard deviation. Otherwise, the data will be summarised using the median and interquartile range. We will not undertake inferential statistics for baseline characteristics but will note the clinical importance of any imbalance.

#### Analysis for the Primary endpoint

We will compute the posterior probability that EpiDex results in a lower probability of hospitalisation than placebo using hierarchical Bayesian relative risk regression, adjusted for site. The prior for the treatment effect is from our expert elicitation exercise [17]. We will report the mean and 95% high-density posterior credible interval for the absolute risk difference and relative risk across the two treatment groups. A secondary analysis of the primary outcome will include only those participants who present with their first episode of bronchiolitis and an analysis that assumes all participants with missing outcome data were not admitted to hospital. All models will include minimally informative priors for the variance parameters [24] and other coefficients [25].

### Analysis for Secondary endpoints

#### Secondary Outcomes

Secondary outcomes one, two and three will be analysed using relative risk regression, like the primary outcome, with minimally informative priors for all parameters. We will report the probability that EpiDex is superior to placebo, the mean absolute risk difference and relative risk and 95% credible intervals. For secondary outcome two, all cause admission up to 21 days after enrolment, we will undertake a secondary analysis that assumes all participants with missing outcome data were not admitted to hospital. For secondary outcome four, costs related to health care expenditure, we will undertake a separate formal economic analysis.

#### Safety Outcomes

Safety outcomes one, two and three will be analysed using relative risk regression with minimally informative priors [26]. We will report the mean absolute difference, the 95% credible interval and the probability of increased chance of adverse events for EpiDex. The number and percentage of observed events for each of the four safety outcomes will also be reported.

Adverse events will be coded using Medical Dictionary for Regulatory Activities (MedDRA) [27] and counted at most once for each participant. We will also present the severity and frequency of the adverse event by System Organ Class.

#### Exploratory Outcomes

Exploratory outcomes (a) – (d) will be analysed using relative risk regression and report the posterior mean absolute-risk difference and relative risk with corresponding 95% credible intervals. Exploratory outcomes (e) and (f) are uncensored time-to-event outcomes and will be analysed as a Bayesian Cox proportional hazards model fit with the Integrated Nested Laplace Approximation [28] method in R [29], adjusted for site. We will report median hazard ratios and their corresponding 95% credible intervals.

Exploratory outcome (g) is a repeatedly measured binary outcome and will be analysed using a hierarchical Bayesian relative risk regression to adjust for patient. From this model, we will report mean relative risks alongside 95% credible intervals. Exploratory outcome (h) will initially be analysed using a Bayesian linear model to report the mean difference in expenses alongside a 95% credible interval. Graphical methods will be used to assess normality of residuals and a lognormal or gamma model will be fit, if required. Exploratory outcomes (i) and (j) will not be analysed at the primary trial analysis of the trial. Thus, a separate analysis plan will be developed before the completion of the 18-year follow-up period.

#### Subgroup analyses

We will perform six planned subgroup analyses to estimate the subgroup-specific treatment effect for the primary outcome. These subgroups are:

1. patients aged <6 months and ≥6 months.
2. patients who developed the first respiratory symptoms, including coryza and cough or fever, within 48 hours of enrolment,
3. patients with a personal history of eczema,
4. patients with parents and/or siblings who have a history of atopy,
5. patients stratified by northern and southern hemispheres, and
6. patients stratified by receipt of nebulised or MDI-s medications: yes/no

For these subgroup analyses, we will perform relative risk regression with main effects for subgroup assignment and treatment group and an interaction between treatment and subgroup. Positive or negative subgroup effects will be declared if the interaction effect has a 99% probability of being greater or less than 0, respectively. We expect the treatment will be more effective in those participants with onset of illness within 48 hours, a personal history of eczema, and a family history of atopy. We do not anticipate differences due to hemisphere of participating hospital or method of inhalation.

#### Additional analyses

If the analyses are adjusted for missingness, we will also perform a complete case analysis by excluding subjects with missing values from the analysis to understand the sensitivity of the conclusions to missing data. We will also undertake a prior sensitivity analysis for the primary analysis using a minimally informative prior for the treatment effect.

#### Heterogeneity of treatment effects analysis

Due to the heterogeneity of bronchiolitis, we will also perform a heterogeneity of treatment effect analysis using a predictive approach [30, 31]. We will use a risk modelling approach by developing a multivariable risk model adjusted for, at a minimum, age, sex, prematurity, length of illness at time of enrolment, history of previous wheezing, personal history of eczema, family history of atopy, household smoke exposure, prenatal smoke exposure, breastfeeding and virus type [32, 33] but ignoring treatment assignment. We will create ten risk strata based on this model and report the median treatment effect with 95% credible intervals within each strata using Bayesian risk regression.

#### Trial Status

The BIPED study was registered on June 25, 2018 and started recruitment in December 2018. Recruitment and follow-up was completed in January 2025. Final monitoring visits will be completed by April 2025. The study database will be cleaned and checked for completeness before the data are analysed. Following this process, the database will be locked, and the statistical analysis will be undertaken using the methods specified in this SAP. We anticipate data locking by end of May 2025.

## Data Availability

This manuscript is a statistical analysis plan. No datasets were used to develop this article as analysis was not undertaken. Thus, this consideration is not applicable.

## List of Abbreviations

BIPED: Bronchiolitis in Infants Placebo Versus Dexamethasone Study
ED: emergency department
SAP: statistical analysis plan
RSV: Respiratory Syncytial Virus
PERC: Pediatric Emergency Research Canada
PREDICT: (Paediatric Research in Emergency Departments International Collaboration
PERN: Pediatric Emergency Research Networks
MDI-s: metered dose inhaler and spacer
EpiDex: a combination of inhaled epinephrine and dexamethasone
REB: Research Ethics Boards
DSMB: data and safety monitoring board
DCC: data coordination centre
WCHRI: Women and children’s Health Research Institute
REDCap: research electronic data capture
ICU: intensive care unit
CPAP: continuous positive airway pressure
ALC: average length criterion
CONSORT: Consolidated Standards of Reporting Trials
RDAI: respiratory distress assessment index
MedRA: Medical dictionary for regulatory activities.

## Declarations

### Ethics approval and consent to participate

REB approval has been obtained for all study sites In Canada, approval has been obtained from Sainte-Justine’s UHC Research Ethics Committee (Study ID 2019-2062), the Children’s Hospital of Eastern Ontario REB (CHEO REB protocol 18/07 CTO and Lawson Approval R-18-669), the University of Manitoba Biomedical Research Board (HS22184[B2018:09]), the Health REB - Biomedical Panel (Edmonton) (Study ID Pro00083863), and the Conjoint Health REB, University of Calgary (Ethics ID REB18--0424). In New Zealand approval has been granted by the Health and Disability Ethics Committee (Ethics ID 19/STH/210). In Australia approval has been granted by the Child and Adolescent Health Services Human Research Ethics Committee (RGS3033). Consent for enrollment will be obtained from the legal guardians of eligible infants.

### Consent for publication

Not applicable

### Availability of data and material

No datasets were used to develop this article as analysis was not undertaken. Thus, this consideration is not applicable.

### Authors’ Contributions

AH, DR, TR, KV, NW, EO, MO, PP, TPK, SRD and ACP were involved in the conception and design of the BIPED study and the SAP. AH and DR drafted the manuscript. TR, KV, EO, MO, PP, TPK, SRD and ACP offered substantive revisions. All authors read, edited and approved the final manuscript. All individuals mentioned in the Acknowledgements are members of the BIPED Study Group.

## Acknowledgments

We thank all members of the KidsCAN PERC Innovative Pediatric Clinical Trials BIPED Study Group and the KidsCAN PERC Innovative Pediatric Clinical Trials Methods Core (Eleanor Pullenayegum, Jeff Round, and Andy Willan) for assistance in developing the protocol and statistical analysis plan for the BIPED trial. The BIPED study would also like to acknowledge DSMB members Dr. Garth Meckler, Dr. Mark Roback, Dr. Anupam Kharbanda, Dr. Eyal Cohen and Dr. Lise Nigrovic and members of KidsCAN PERC Innovative Pediatric Clinical Trials. We want to thank the Pediatric Emergency Research Canada (PERC) and PREDICT network of health care professionals and the KidsCAN Trials Network for their contribution and support to this project and pediatric clinical research in Canada, Australia and New Zealand. Finally, we thank Jingxian (Phebe) Lan for leading the expert elicitation exercise to determine the prior distributions used in the primary analysis.

The BIPED Study group comprises: Sharon O’Brien (Perth Children’s Hospital, Nedlands, Western Australia, Australia), Meredith L Borland (Perth Children’s Hospital, Nedlands, Australia), David W Johnson (University of Calgary, Calgary, Alberta, Canada), Joseph J Zorc (Children’s Hospital of Philadelphia, Pennsylvania, USA), Suzanne Schuh (Hospital for Sick Children, Toronto, Ontario), Medhawani Rao (Starship Children’s Hospital, Auckland, New Zealand), Megan Bonisch (Starship Children’s Hospital, Auckland, New Zealand), Simon S Craig, Monash University, Melbourne, Victoria, Australia), Serge Gouin (Le Centre Hospitalier Universitaire Sainte-Justine, Montréal, Québec, Canada), Amit Kochar (Women and Children’s Hospital, Adelaide, South Australia, Australia), Graham C Thompson (University of Calgary, Calgary, Alberta, Canada), Chris Lash (Middle Hospital, Auckland, New Zealand), Alexandra Wallace (Waikato Hospital, Hamilton, New Zealand), Andrew Dixon (Stollery Children’s Hospital, Edmonton, Alberta, Canada), Scott Sawyer (HSC Children’s Hospital, Winnipeg, Manitoba, Canada), Gary Joubert (Children’s Hospital London Health Sciences Centre, London, Ontario, Canada)

